# Ambient AI Documentation in Mixed-Language Encounters: A Heuristic Evaluation of Reenacted Mandarin–English and Spanish–English Clinical Conversations

**DOI:** 10.64898/2026.05.19.26353603

**Authors:** Di Hu, Daniel Flores, Lidia Flores, Ruby Chien, Kyle Lam, Emilie Chow, Yawen Guo, Steven Tam, Danielle Perret, Deepti Pandita, Kai Zheng

## Abstract

Ambient AI documentation systems rely on automatic speech recognition to transcribe patient–provider conversations before generating clinical notes. However, little evidence exists on how these systems perform in mixed-language clinical encounters. We conducted a mixed-methods heuristic evaluation of an ambient AI documentation tool using 24 reenacted primary care conversations, including 12 Mandarin–English conversations developed from real-world encounter excerpts and 12 Spanish–English adapted counterparts. Quantitative analyses measured mixed error rate (MER) and code-switching. Overall MER was low, with a median of 4% and less variation in Spanish–English conversations, and 9% in Mandarin–English conversations, but with outliers reaching 67%. The system generally detected language switches reliably, although deletions occurred frequently in Mandarin–English transcripts at switch points. Qualitative analysis revealed transcription errors related to phonetic similarity, automatic translation, clinical terminology recognition, and language-specific challenges. These findings highlight considerations for improving ambient AI tools to support multilingual providers in delivering care for linguistically diverse populations.

## Introduction

Ambient artificial intelligence (AI) clinical documentation tools are increasingly being deployed across major U.S. health systems to address documentation burden and clinician burnout^1–5^ . These tools capture audio from patient–provider conversations, apply automated speech recognition (ASR) technology to generate transcripts, and then use generative AI to draft clinical notes based on those transcripts. Early evaluations in real-world clinical settings suggest that ambient AI may improve documentation efficiency and clinician experience, including reduced time spent on note writing^1–3^, lower perceived burnout^1,2,4,5^, and improved job satisfaction^2,4^. Together, these findings indicate that ambient AI has the potential to alleviate documentation-related burden and support clinician well-being.

As these tools move into routine clinical practice, they must operate within the linguistic realities of U.S. healthcare. The U.S. has the largest immigrant population in the world^6^, with approximately 53.3 million foreign-born residents^7^, nearly half of whom report limited English proficiency (LEP)^8^. Language barriers are known obstacles to effective clinical communication and are associated with suboptimal patient-clinician interactions, and increased risk of adverse outcomes^9,10^. Patients with LEP may prefer to communicate in or switch to their primary language during clinical encounters, particularly when discussing complex health information^11^, and care from language-concordant clinicians has been shown to improve care quality and patient experience^9,10,12^. In practice, navigating language support while delivering care may impose additional, often uncompensated workload on clinicians, which may also contribute to burnout^13^. Indeed, clinicians have particularly noted in previous studies that multilingual capabilities in ambient AI documentation tools are valuable and should be improved for caring linguistically diverse patient populations^14,15^. Ambient AI systems should therefore support this need so that clinicians serving these patients can receive comparable benefits.

However, prior research has shown that ASR performance generally declines in mixed-language conditions compared with monolingual speech^16^. In multilingual encounters, patients and clinicians may alternate between languages within the same conversation. This phenomenon, known as code-switching (CS), occurs as speakers adapt their language to improve comprehension or achieve communication goals during consultations^17,18^. CS has long been recognized as a challenge for ASR, especially for low-resource languages with limited available speech data for model training and evaluation^16,19^. In the context of ambient AI documentation, errors introduced during transcription may propagate into AI-generated note drafts, potentially increasing clinician review effort and reducing trust in the system^20^. Despite the need for multilingual support and the recognized technical challenges, no existing Ambient AI evaluations have focused on multilingual encounters, leaving little empirical evidence on how these systems perform in mixed-language clinical conversations.

To address this gap, we conducted a mixed-methods heuristic evaluation of an ambient AI documentation tool, Abridge, in multilingual clinical conversations. As an exploratory step, this study examines ASR performance in transcribing mixed-language primary care conversations in which both patients and clinicians engage in CS. For evaluation, we reenacted 12 Mandarin–English patient–provider conversations developed from real-world primary care encounter excerpts and 12 Spanish–English counterparts translated and adapted from the Mandarin–English reference transcripts. These language pairs were selected for this preliminary heuristic exploration because they are among the most common non-English languages represented in UCI Health’s patient population and because the study team included bilingual researchers and clinicians with the language expertise needed to construct, reenact, and evaluate these conversations. This design enabled an initial comparison of ASR performance across language pairs while holding clinical content and encounter structure relatively constant. Our study aims to (1) assess ASR performance in mixed-language clinical conversations, (2) identify common transcription errors that may be clinically relevant, increase clinician review effort, or affect note quality, and (3) highlight considerations for future Ambient AI evaluation and improvement to better support clinicians serving LEP and linguistically diverse patient populations. By focusing on mixed-language conversations based on real-world encounters, this study provides practice-grounded evidence to inform the future design and evaluation of ambient AI documentation tools that are more reliable and equitable in multilingual clinical care.

## Methods

### Study Setting and Dataset Construction

This study evaluated the performance of an ambient AI documentation tool (Abridge, Abridge AI, Inc., Pittsburgh, PA, USA) implemented at UCI Health using reenacted clinical conversations involving mixed-language communication. Abridge uses its proprietary ASR to process audio and currently supports transcription in 28 languages, generating a fully English note at the end^21^. To construct the evaluation dataset, we first developed 12 Mandarin–English reference transcripts based on selected conversation excerpts from 15 real-world primary care encounters involving Mandarin-speaking patients who spoke English as a second language and two bilingual physicians (EC, RC). The excerpts were selected through review of existing de-identified audio recordings from the ambient AI tool. Each excerpt included segments in which both languages were spoken, and the clinical context was sufficiently complete to function as a standalone interaction. The clinical context and language use in the conversations, particularly how patients and providers switched between languages, were preserved to reflect realistic mixed-language communication patterns in clinical practice. To simplify the conversational structure for evaluation purposes, excerpts involving additional participants such as caregivers were removed or rewritten to maintain a two-speaker interaction. This study was deemed Non-Human Subjects Research and registered with the Institutional Review Board at the University of California, Irvine.

The developed Mandarin–English reference transcripts were reviewed by a resident physician (RC) and an informatics researcher (DH), both bilingual in Mandarin and English, with minimal edits to ensure accurate clinical content and clear semantic meaning. The transcripts were then translated into Spanish–English dialogue and reviewed by an informatics researcher (LF) and a medical student (DF), both bilingual in Spanish and English, to preserve clinical meaning and ensure that language expressions and CS patterns were representative of Spanish-English communication styles. The Spanish–English transcripts were designed as adapted parallel scenarios rather than independently developed from Spanish–English clinical encounters. This design supported comparison of language-pair differences in ASR behavior by maintaining comparable clinical content, speaker roles, and code-switching opportunities, but introduced a validity tradeoff because adapted Spanish–English phrasing may not fully reflect naturally occurring Spanish–English clinical communication. The finalized transcripts were reenacted and recorded to generate standardized audio inputs for ASR evaluation. During reenactments, the two informatics researchers performed the patient roles, while the resident physician and the medical student performed the provider roles, reading out loud the dialogue directly from the reference transcripts. Recordings were conducted in a private meeting room using a mobile phone microphone (iPhone 16 Pro, Apple, Inc., Cupertino, CA, USA) through the recording function in the Abridge application, replicating the workflow used in routine clinical deployment at UCI Health. This approach standardized recording conditions, including speaker positioning and background noise, while preserving the conversational structure and language mixing defined in the transcripts.

### Mixed-Methods Design

We used a sequential explanatory mixed-methods design guided by a pragmatic paradigm. Quantitative ASR metrics were first calculated to describe transcription accuracy, code-switching detection, and error distributions across the two language-pair datasets. Qualitative descriptive error analysis was then conducted to examine the content and context of the identified errors and explain patterns observed in the quantitative results, including differences in MER, deletion errors, and code-switching performance. Given the exploratory and heuristic purpose of this study, this design allowed both quantification of transcription performance and examination of what kinds of errors occurred, when they appeared, and how they could affect clinical documentation.

### Quantitative Analysis of ASR Performance

We conducted quantitative analyses to evaluate the ASR performance of the ambient AI documentation system. Transcripts were tokenized and normalized (e.g., lowercasing and punctuation removal) prior to any computations. Reference and ASR transcripts were aligned at the token level for comparison. We summarized median, range, 25th percentile, and 75th percentile as descriptive statistics for audio duration, transcript length, and the proportion of each language used within the conversation. Errors were quantified as substitutions (S), deletions (D), and insertions (I) of words or characters in the ASR transcripts. Mixed Error Rate (MER), combining word error rate (WER) and character error rate (CER), was computed as (S + D + I) / N, where N is the number of tokens in the reference transcript, treating tokens from both languages as a single sequence. We also operationalized CS recall, precision, and F1, to examine how well the system captured language switching during mixed-language conversations. For each excerpt, we identified points where the spoken language switched between English and Spanish or Mandarin and checked if the code-switching points in the reference were correctly identified in the hypothesis transcripts. For Spanish-English cases, when some words (e.g., “a”, “he,” “son,” “uses”) appear in both languages and cannot be reliably distinguished as English or Spanish by algorithms in mixed-language written text, researchers manually labeled these tokens based on the audio recordings. All analyses were conducted using Python 3.12.2.

### Qualitative Descriptive Error Analysis of ASR in Mixed-Language Clinical Transcription

To further understand specific ASR challenges in mixed-language clinical conversations, we conducted a qualitative descriptive analysis of transcription errors. This analysis focused on errors identified through the quantitative token-level alignment, including substitutions, deletions, and insertions, as well as transcripts with high MER, large deletion counts, or low code-switching recall. Because this analysis focused on ASR error classification, we aimed to identify recurring error categories across transcripts rather than assess thematic saturation. The same bilingual researchers who reenacted the conversations (DH, RC, DF, LF) reviewed the AI-generated transcripts alongside the reference transcripts for each excerpt, with S/D/I errors flagged through quantitative alignment. For each language pair, the first author (DH) and at least one bilingual reviewer (RC, DF, or LF) first reviewed three transcripts to inductively develop an initial error codebook. Reviewers examined each error in its surrounding conversational context, with particular attention to code-switching, medical terminology, and potential clinical relevance. The team met to compare codes, discuss disagreements, resolve conflicts through consensus, and refine the codebook. After the initial codebook was developed, DH applied the refined codes to all ASR errors across the remaining transcripts. Recurring codes were then reviewed and merged into broader error categories within and across language pairs. These qualitative findings were used to help explain and contextualize the ASR performance patterns observed in the quantitative analysis.

## Results

### Data Overview

We present descriptive statistics for our dataset and the results of the ASR performance metrics in Table 1. A total of 24 reenacted mixed-language clinical conversation recordings were analyzed, including 12 in Spanish–English and 12 in Mandarin–English. Because the Spanish–English conversations were adapted as parallel scenarios from the Mandarin–English reference transcripts, the two language-pair datasets involved comparable clinical content and encounter structure. Despite this shared content, Mandarin–English conversations were generally longer and produced longer transcripts than the adapted Spanish–English conversations, as reflected in higher median audio duration and transcript token counts. In the Spanish–English excerpts, the reference transcripts and ASR outputs contained very similar token counts. In contrast, the Mandarin–English ASR transcripts were noticeably shorter than their reference transcripts, indicating substantial deletions. The distribution of languages was comparable across groups. The conversations consisted of approximately a 35–65 mixture of the two languages, with non-English tokens constituting the majority of speech.

**Table 1.** Mixed-language clinical conversation characteristics and ASR performance metrics.

| Metrics | Spanish–English (n=12) |  | Mandarin–English (n=12) |  |
| --- | --- | --- | --- | --- |
|  | Median (Q1–Q3) | Range | Median | Range |
| **Conversation Characteristics** |  |  |  |  |
| Audio Duration (seconds) | 192.50<br>(163.75–245.25) | 112–362 | 204.00<br>(166.75–244.25) | 130–378 |
| Reference Transcript Length (tokens*) | 413.00<br>(360.75–515.50) | 239–794 | 541.50<br>(475.25–637.75) | 317–1040 |
| ASR Transcript Length (tokens*) | 412.50<br>(357.25–508.00) | 241–770 | 525.50<br>(334.75–579.75) | 173–958 |
| English in Reference Transcript (%) | 35.75<br>(23.19–41.03) | 7.20–65.72 | 32.05<br>(20.24–36.25) | 3.02–57.70 |
| English in ASR Transcript (%) | 36.01<br>(22.24–40.11) | 5.93–65.51 | 26.50<br>(16.25–32.78) | 3.07–65.32 |
| Non-English in Reference Transcript (%) | 64.25<br>(58.97–76.81) | 34.28–92.80 | 67.95<br>(63.75–79.76) | 42.30–96.98 |
| Non-English in ASR Transcript (%) | 63.99<br>(59.89–77.76) | 34.49–94.07 | 73.50<br>(67.22–83.75) | 34.68–96.93 |
| **ASR Error Metrics** |  |  |  |  |
| Substitutions (n) | 9.00<br>(4.75–11.25) | 1–19 | 11.50<br>(9.25–22.00) | 5–47 |
| Deletions (n) | 3.00<br>(2.00–10.00) | 0–26 | 48.00<br>(6.00–113.50) | 4–247 |
| Insertions (n) | 2.00<br>(1.00–3.00) | 0–4 | 3.50<br>(1.00–6.00) | 0–15 |
| Mixed Error Rate (%) | 3.81<br>(2.71–4.47) | 1.26–8.98 | 9.17<br>(4.63–20.11) | 2.26–67.10 |
| **Code-switching Metrics** |  |  |  |  |
| Code-Switches in Reference Transcript (n) | 67.00<br>(55.25–89.25) | 34–143 | 60.00<br>(45.25–84.75) | 20–141 |
| Code-Switches in ASR Transcript (n) | 60.50<br>(52.75–77.75) | 32–128 | 46.00<br>(27.75–77.75) | 12–110 |
| Correctly Detected Code-Switches (n) | 58.00<br>(51.25–77.25) | 30–124 | 46.00<br>(27.25–74.25) | 11–103 |
| Code-Switch Detection Recall | 0.88<br>(0.82–0.95) | 0.76–1.00 | 0.86<br>(0.71–0.92) | 0.24–1.00 |
| Code-Switch Detection Precision | 0.97<br>(0.95–0.99) | 0.89–1.00 | 0.98<br>(0.93–1.00) | 0.92–1.00 |
| Code-Switch Detection F1 | 0.92<br>(0.88–0.97) | 0.83–1.00 | 0.92<br>(0.81–0.94) | 0.38–1.00 |

\* Tokens refer to English words and Chinese characters treated as individual units during evaluation.
Q1: 25th percentile, Q3: 75th percentile.

### ASR Performance Metrics

ASR transcription accuracy was high across both evaluated language-pair datasets, but the adapted Spanish–English conversations showed a noticeable advantage, with smaller numbers of substitutions, deletions, and insertions, and a low median MER of about 4%. Performance was also consistent across excerpts. In contrast, Mandarin–English conversations had more transcription errors, with a median MER of about 9% and considerably greater variability, including a much wider interquartile range (Q1–Q3: 4.63–20.11) with a highest error rate of 67%. While the median substitution and insertion errors were comparable to those in Spanish–English conversations, deletion errors (Median: 48.00) were the most pronounced error type in Mandarin–English mixed conversations. These deletion errors contributed substantially to the observed performance differences, with five ASR transcripts deleting more than 10% of their total tokens.

Spanish–English conversations contained slightly more code-switches in the reference transcripts (median = 67.00) than Mandarin–English conversations (median = 60.00). Despite these differences in token-level recognition accuracy, the system performed generally well in identifying code switches for both mixes. For Spanish–English conversations, the system achieved high recall and precision, resulting in a strong median F1 score. Mandarin–English conversations showed a similar median F1 score, although recall varied more widely across excerpts, with the lowest at 0.24. This lower recall may be related to deletion errors in the ASR transcripts, which could lead to missed CS tokens and reduce the number of correctly detected switches. The qualitative analysis below further examines how deletion errors and local transcription instability appeared around some code-switching points.

### Challenges for ASR for Mixed-Languages Clinical Transcription

To explain the quantitative performance patterns and identify common ASR challenges in mixed-language clinical conversations, we conducted a qualitative review of ASR transcription errors in context. This review showed that several ASR limitations in this setting arose not only from clinical context and code-switching separately, but also from their interaction. The qualitative findings also revealed that favorable aggregate metrics could still coexist with clinically relevant mistranscriptions and instability around code-switching points. We identified four recurring error patterns, including errors due to phonetic similarity across languages, automatic language translation, difficulty recognizing clinical terminology, and language-specific challenges. For each error type, clinically relevant examples were selected from transcripts and presented to illustrate how these errors may appear in practice and affect clinical documentation.

#### Errors Due to Phonetic Similarity Within and Across Languages

A common error pattern in both datasets involved substitutions driven by phonetic similarity, where the ASR output approximated the spoken token in sound but produced a different word. One type of such error involved words, characters, or phrases with **similar pronunciation but different meanings within the same language**, a pattern that also commonly occurs in monolingual speech recognition. For example, in a Mandarin conversation segment, “*肝 胆*” (*gān dăn*, liver and gallbladder) was transcribed as “*肝 脏*” (*gān zàng*, liver) when the patient requested a referral. Similarly, “*摔 跤*” (*shuāi jiāo*, fall) appeared as “*睡 觉*” (*shuì jiào*, sleep) when a patient described the cause of a symptom, and “*刺 刺*” (*cì cì*, tingling) was transcribed as “*次 次*” (*cì cì*, every time) when the patient described their sensation. In the Spanish examples, “*mi presión*” (my blood pressure) was transcribed as “*depresión*” (depression) and “*amarilla*” (yellow), used to describe the color of mucus, was transcribed as “*amaría*” (would love). Although aggregate MER was low overall, these mistranscriptions could misrepresent clinically relevant information from the conversation in the transcript and subsequent note draft.

Another pattern was observed when terms with **similar pronunciation carried different meanings across languages**, which is specific to mixed-language conversations. For example, the English word “*bone*” was transcribed as “*泵*” (*bèng*, pump), a Chinese character with identical pronunciation. Although both terms can appear in clinical contexts, they have very different meanings. Similarly, the Mandarin phrase “*深 呼 吸*” (*shēn hū xī*, take a deep breath), commonly spoken during a physical exam, appeared as “*some who she*,” where each Mandarin character was mapped to a phonetically similar English word, producing a meaningless output. Cross-language substitutions also occurred in Spanish–English cases, “*Tos”* (cough) was transcribed as “those,” “*blood test*” as “*brotes*” (sprouts), and “*viste al*” (did you see the) as “*we still have*.” These examples illustrate how phonetic overlap across languages can lead the ASR system to generate incorrect and contextually inappropriate tokens.

#### Errors Due to Automatic Language Translation

Another recurring discrepancy between the reference and ASR transcripts was due to automatic language translation, in which the ASR output appeared to **translate or normalize speech rather than transcribe it as spoken**. This pattern was observed more frequently in the adapted Spanish–English dataset despite its lower MER and fewer deletion errors, suggesting that stronger aggregate accuracy can still coexist with transcript fidelity issues in how spoken language is evaluated. Although the pronunciations of some English and Spanish words are not identical, the English words “*carbohydrates*,” “*creatinine*,” and “*chemotherapy*” spoken in the audio were transcribed as their Spanish equivalents “*carbohidratos*,” “*creatinina*,” and “*quimioterapia*.” These Spanish words may be similar enough in pronunciation for an ASR system that processes multiple languages to recognize them in either language based on the surrounding CS context. However, a similar phenomenon was also observed in the Mandarin–English dataset. Unlike the Spanish examples, Mandarin and English words with the same meaning rarely share similar pronunciations, suggesting that automatic language translation substitutions occur independently of phonetic similarity.

In several cases, English speech was transcribed as Chinese characters or rendered in Pinyin (the official romanization system for Mandarin Chinese). For instance, the English word “*happen*” was transcribed as “*发 生*” (*fā shēng*, happen), possibly because this single-word CS appeared within a relatively long Mandarin segment, and the Mandarin character “*胸*” (*xiōng*, chest) appeared as the Pinyin “*xiong*.” This behavior was not limited to individual words or short phrases; in some cases, entire sentences were translated during transcription. Although automatic language translation may appear less harmful when the meaning is preserved, we observed that the translations were not always accurate. For example, the ASR output produced “neurologista” for “neurologist,” which is not a valid Spanish word. In the Mandarin–English dataset, the system translated “wrist” as “*肘*” (*zhǒu*, elbow) and “arm” as “*腕*” (*wàn*, wrist), leading to confusion about the body parts being discussed. In another case, “*胃 镜*” (*wèi jìng*, gastroscopy) was rendered as the pseudo-Pinyin “*weiji*” instead of the correct Pinyin “*weijing*,” producing an output that was difficult to interpret later in the transcript. Even when the translation itself was correct, language translation introduced alignment challenges between the reference and ASR transcripts. Equivalent meanings may be represented with different token lengths across languages, shifting token positions between transcripts for evaluation. For example, the number “*veinticinco*” is a single word in Spanish, two words in English (“*twenty five*”), and three characters in Chinese (“二 十 五”). These observations suggest that when ASR systems prioritize linguistic normalization over faithful transcription, the resulting outputs may alter the language of the original speech and introduce additional challenges for transcript interpretation and evaluation.

#### Errors in Recognizing Clinical Terminology

Errors in recognizing clinical terminology appeared frequently in the ASR transcripts, illustrating how clinically meaningful errors could occur even when overall ASR performance was favorable. These included **non-recognized or misspelled laboratory tests, medication names, medical devices, and condition terms**. Because care was delivered in the U.S. healthcare system, standardized medical terminology, particularly medication brand names, was often retained in English even when the surrounding conversation occurred in another language. However, many medical terms have uncommon pronunciation and spelling patterns, which can make them difficult for ASR systems to recognize accurately. Examples observed in the Mandarin–English transcripts include medication and device names that were not recognized or were transcribed incorrectly, such as “*leqvio*” as “*lekvia*,” “*amlodipine*” as “*enlodipine*,” and “*zio patch*” as “*xylem patch*.” These errors often occurred when patients (played by informatics researchers without a medical education background) pronounced the terms themselves, suggesting that familiarity with the terminology and pronunciation may influence ASR recognition.

In the Spanish–English conversations, another pattern suggested that the **ASR system tended to favor more frequently used words in the surrounding language rather than recognizing the intended medical terminology**. This occurred when medication or laboratory names contained segments whose pronunciation resembled common Spanish words, confusing the ASR system. For example, *losartan* was transcribed as “*los zartan*,” where “*los*” was interpreted as the Spanish article and separated from the rest of the medication name. Similarly, *cystatin C* appeared as “cistatin sí,” because the letter “*C*” was interpreted as *sí* (yes) in Spanish. While part of the term was recognized as Spanish, the remaining segments often appeared as misspelled or phonetically similar words, consistent with clinical terminology errors observed in the Mandarin cases. This type of error reflects the interaction between clinical terminology recognition and multilingual processing challenges.

#### Language-Specific Challenges

While the error patterns identified appeared across both language groups, findings also revealed distinct language-specific challenges. In the **Spanish–English** conversations, some errors involved grammatical or tense variations within Spanish words, such as “*fumar*” → “*fumo*” (infinitive “to smoke” → first-person present “I smoke”) or “*terminé*” → “*terminó*” (first-person past “I finished” → third-person past “he/she finished”). These substitutions changed the grammatical form of the original word and reduced the textual clarity of the transcript. Another error pattern specific to Spanish–English conversations involved substitutions in which the ASR output shifted languages while preserving the intended meaning. For example, the English word “*is*” was transcribed as “*es*”, “*stable*” as “*estable*”, and “*in*” as “*en*”. Because these substitutions reflect both phonetic similarity and semantic equivalence, it can be difficult to determine the intended language of the word during transcription and to distinguish whether the substitution results from language detection errors or automatic language translation during evaluation. Although these substitutions appear less likely to affect interpretation of the clinical content compared with medical terminology errors, their actual impact remains unclear.

In the **Mandarin–English** conversations, unique error patterns also emerged that helped explain the higher deletion counts and wider MER variability observed in the quantitative results. Six of the twelve ASR outputs exhibited large text drops (deletion exceeding 50 tokens), including cases occurring at or near CS points. Insertions of garbled letter sequences and repeated or looping characters were also observed around CS points only in the Mandarin–English ASR transcripts. These patterns suggest that language transitions may represent vulnerable moments for Mandarin transcription stability. The Mandarin outputs also contained errors involving Chinese pronouns. Because the pronouns for “*he*,” “*she*,” and “*it*” share the same pronunciation in Mandarin, the characters “*他*” (*tā*, he), “*她*” (*tā*, she), and “*它*” (*tā*, it) may be used interchangeably in transcription. In several cases, the ASR system produced the wrong character even when contextual cues from surrounding English words indicated the intended referent. Another Mandarin-specific issue relates to the coexistence of two written forms of Chinese characters: *Simplified* and *Traditional*. Although they represent the same spoken Mandarin words, these two writing systems differ in character structure. The ASR system occasionally switched between *Simplified* and *Traditional* Chinese characters within the same transcript, resulting in inconsistent written outputs.

## Discussion

Our evaluation shows that MER across the evaluated examples was low overall, with median values below 10% for both the Mandarin–English and adapted Spanish–English conversation excerpts. These MER values were lower than Abridge’s general benchmark^22^, which may be explained by the selective nature of our dataset, as it consisted of relatively short excerpts with routine clinical context and a small sample size. In addition, the ASR system performed well in detecting code-switching in our datasets. However, a clear performance difference was observed between the two language groups, with ASR performing better and more robustly in Spanish–English conversations, while performance in Mandarin–English excerpts was less stable, with some excerpts being considerably more error-prone. This difference was largely driven by deletion errors, including large text drops in several Mandarin–English transcripts occurring at or near code-switching points. It may also reflect the lower prevalence of Mandarin relative to Spanish in many U.S.-based ASR training datasets and structural differences between the languages. Despite the generally favorable aggregate results, transcription errors remained in the ASR outputs used for clinical documentation, and some could have clinical implications if not detected during clinician review. This suggests that ASR metrics alone may underestimate clinically relevant transcription risks in mixed-language encounters. By combining quantitative evaluation with qualitative analysis, this study provides an initial but detailed understanding of ASR performance in mixed-language clinical interactions and the types of errors that arise in this setting, highlighting recognition challenges and domain-specific issues related to clinical terminology and multilingual conversational context.

Many of the challenges identified in this study are consistent with findings from the existing literature on ASR for CS. Previous studies have shown that ASR systems often struggle to recognize CS in mixed-language speech, where phonetic boundaries between languages can blur and language segments may be misidentified^16,23,24^. At the same time, we identified error patterns that are more language-specific and have received less attention. For example, errors such as the alternating generation of *Traditional* versus *Simplified* Chinese characters within a single transcript. This inconsistency could be addressed by allowing providers to set the desired output format for the transcript. Also, some phrases were transcribed as Pinyin or pseudo-Pinyin instead of Chinese characters, which may occur when the ASR system cannot confidently match a Chinese character or phrase to the spoken pronunciation. This behavior may be useful when discussing culturally specific concepts whose English usage is commonly represented in Pinyin, such as “*Tai chi*” or certain terms in Chinese medicine. However, the Pinyin spelling still needs to be accurate for it to be meaningful. Another unexpected observation is the automatic translation of words, phrases, or sentences spoken in English into the other language, which occurred more frequently in the Spanish–English conversations. In these cases, the translation can cause misalignment between the ASR output and the reference transcript. Although these variations typically do not substantially change the semantic meaning of the sentences, they introduce unnecessary randomness and inconsistencies in the transcription outputs for downstream note generation. If not recognized and properly handled, such variations may also skew metric calculations for system performance. These observations suggest the need to account for such patterns when evaluating ASR systems and developing evaluation approaches tailored to mixed-language clinical conversations.

Additionally, our findings provide further insight into how challenges in recognizing language CS manifest in clinical conversations. These issues may be more pronounced in clinical applications. Real-world patient–provider conversation data are difficult to collect due to privacy constraints^25,26^, and the availability of multilingual CS data is also limited^16,24^. As a result, data representing multilingual clinical interactions may be even harder to obtain. Efforts should be dedicated to construct such training data to improve ASR. The potential impact of transcription errors may also be greater in clinical contexts. In several examples from our data, both the intended and the substituted words are clinically plausible but carry very different meanings. Clinical contexts introduce additional complexity because conversations often include specialized terminology that can be difficult for ASR systems to recognize accurately. Prior work on speech recognition for clinical documentation has noted similar challenges related to domain-specific vocabulary^27,28^. Our findings suggest that these difficulties may be amplified when medical terms with uncommon English pronunciations are embedded in speech that alternates between languages where similar pronunciations correspond to more prevalent words, and when pronunciation and accent vary across speakers. Large text drops and repeated text loops at CS points may cause clinically relevant information to be missed entirely and affect the final generated note. Therefore, further systematic and repeated testing is needed to understand when and how CS triggers large deletions or text repetition and to determine whether confusion from mixed-language input affects the backend model processing. Addressing these challenges is important to avoid increasing the review effort for clinicians who serve multilingual and LEP patients for AI-assisted documentation, reducing the expected potential of multilingual AI documentation for burnout reduction. Before such errors can be fully prevented, simple quality signals or alters around code-switching points could support clinician review.

This study has several limitations. The dataset is relatively small and consists of selected excerpts rather than full clinical encounters. In addition, the audio recordings were reenacted in a controlled environment, which may not fully capture the complexity present in real clinical settings, such as background noise, interruptions, overlapping speech, speaker movement, variation in microphone distance, or room setup. The results should therefore be interpreted as performance under relatively ideal conditions and may represent a lower bound of the errors that could occur in practice. The study also focused on only two language pairs, Mandarin–English and Spanish–English, while many other languages are used in clinical encounters in diverse healthcare settings. The Spanish–English reference transcripts were translated and adapted from transcripts originally developed from Mandarin–English conversations; this parallel-scenario design supported comparison across language pairs by maintaining similar clinical content and encounter structure, but may have introduced phrasing patterns, code-switching structures, or interactional features that differ from naturally occurring Spanish–English clinical communication. Lastly, the evaluation only assessed one ambient AI tool deployed at UCI Health, so findings may not generalize well to other tools or settings. Future work should incorporate independently collected real-world mixed-language conversations for each language pair, assess additional commonly used and underrepresented languages, examine the content and conditions of large deletion errors, evaluate environmental and acoustic factors such as recording device and microphone distance, and investigate how ASR errors affect downstream AI-generated note quality, editing burden, and clinician trust.

## Conclusion

This study provides an initial heuristic evaluation of an ambient AI documentation tool in transcribing mixed-language clinical encounters involving Mandarin–English and Spanish–English conversations. While overall transcription and CS accuracy appeared favorable, our findings show that mixed-language speech introduces errors related to phonetic similarity, automatic language translation, clinical terminology recognition, and language-specific barriers. These errors may affect the reliability of transcripts used for clinical documentation and increase clinician review effort. This work highlights important considerations for evaluating and improving ASR systems used in multilingual healthcare environments. Addressing these challenges is important to ensure that ambient AI documentation tools equitably support clinicians and patients in linguistically diverse clinical settings.

## Data Availability

All data produced in the present study are available upon reasonable request to the authors.

## Notes

### Competing Interest Statement

The authors have declared no competing interest.

### Funding Statement

This study did not receive any funding.

### Summary of Updates

Minor revisions made per reviewers comments.

## References

1. Guo Y, Wang J, Hu D, Tam S, Gilman C, Chow E, et al. Evaluating ambient artificial intelligence documentation: effects on work efficiency, documentation burden, and patient-centered care. J Am Med Inform Assoc. 2026 Feb 1;33(2):273–82. doi:10.1093/jamia/ocaf180

2. Albrecht M, Shanks D, Shah T, Hudson T, Thompson J, Filardi T, et al. Enhancing clinical documentation with ambient artificial intelligence: a quality improvement survey assessing clinician perspectives on work burden, burnout, and job satisfaction. JAMIA Open. 2025 Feb 21;8(1):ooaf013. doi:10.1093/jamiaopen/ooaf013 PubMed PMID: 39991073; PubMed Central PMCID: PMC11843214.

3. Ma SP, Liang AS, Shah SJ, Smith M, Jeong Y, Devon-Sand A, et al. Ambient artificial intelligence scribes: utilization and impact on documentation time. J Am Med Inform Assoc JAMIA. 2024 Dec 17;32(2):381–5. doi:10.1093/jamia/ocae304 PubMed PMID: 39688515; PubMed Central PMCID: PMC11756633.

4. You JG, Dbouk RH, Landman A, Ting DY, Dutta S, Wang JC, et al. Ambient Documentation Technology in Clinician Experience of Documentation Burden and Burnout. JAMA Netw Open. 2025 Aug 21;8(8):e2528056. doi:10.1001/jamanetworkopen.2025.28056

5. Shah SJ, Devon-Sand A, Ma SP, Jeong Y, Crowell T, Smith M, et al. Ambient artificial intelligence scribes: physician burnout and perspectives on usability and documentation burden. J Am Med Inform Assoc. 2025 Feb 1;32(2):375–80. doi:10.1093/jamia/ocae295

6. List of sovereign states by immigrant and emigrant population. In: Wikipedia [Internet]. 2026 [cited 2026 Mar 8]. Available from: https://en.wikipedia.org/w/index.php?title=List_of_sovereign_states_by_immigrant_and_emigrant_population&oldid=1339849517

7. Camarota SA, Zeigler K. CIS.org [Internet]. [cited 2026 Mar 8]. Foreign-Born Number and Share of U.S. Population at All-Time Highs in January 2025. Available from: https://cis.org/Report/ForeignBorn-Number-and-Share-US-Population-AllTime-Highs-January-2025

8. migrationpolicy.org [Internet]. [cited 2026 Mar 8]. State Language Data - US. Available from: https://www.migrationpolicy.org/data/state-profiles/state/language/US

9. Wilson E, Chen AH, Grumbach K, Wang F, Fernandez A. Effects of Limited English Proficiency and Physician Language on Health Care Comprehension. J Gen Intern Med. 2005 Sep;20(9):800–6. doi:10.1111/j.1525-1497.2005.0174.x PubMed PMID: 16117746; PubMed Central PMCID: PMC1490205.

10. Schenker Y, Karter AJ, Schillinger D, Warton EM, Adler NE, Moffet HH, et al. The impact of limited English proficiency and physician language concordance on reports of clinical interactions among patients with diabetes: the DISTANCE Study. Patient Educ Couns. 2010 Nov;81(2):222–8. doi:10.1016/j.pec.2010.02.005 PubMed PMID: 20223615; PubMed Central PMCID: PMC2907435.

11. Karliner LS, Napoles-Springer AM, Schillinger D, Bibbins-Domingo K, Pérez-Stable EJ. Identification of Limited English Proficient Patients in Clinical Care. J Gen Intern Med. 2008 Oct;23(10):1555–60. doi:10.1007/s11606-008-0693-y PubMed PMID: 18618200; PubMed Central PMCID: PMC2533382.

12. Diamond L, Izquierdo K, Canfield D, Matsoukas K, Gany F. A Systematic Review of the Impact of Patient–Physician Non-English Language Concordance on Quality of Care and Outcomes. J Gen Intern Med. 2019 Aug;34(8):1591–606. doi:10.1007/s11606-019-04847-5 PubMed PMID: 31147980; PubMed Central PMCID: PMC6667611.

13. Santiago-Delgado Z, Bhardwaj N, Frazier WT, Collazo A, Abara NO, Campbell KM. The Minority Tax: Stories from Family Physicians [Internet]. 2025 Jan 30. doi:10.3122/jabfm.2023.230495R1

14. Shah SJ, Crowell T, Jeong Y, Devon-Sand A, Smith M, Yang B, et al. Physician Perspectives on Ambient AI Scribes. JAMA Netw Open. 2025 Mar 24;8(3):e251904. doi:10.1001/jamanetworkopen.2025.1904

15. Hundal J, Jain M, McCollom J. Ambient Artificial Intelligence in Health Care Documentation: A Review of Tools, Integration, and Clinical Implications. AI Precis Oncol. 2025 Oct 1;2(5):158–62. doi:10.1177/2993091X251384313

16. Mustafa MB, Yusoof MA, Khalaf HK, Rahman Mahmoud Abushariah AA, Kiah MLM, Ting HN, et al. Code-Switching in Automatic Speech Recognition: The Issues and Future Directions. Appl Sci. 2022 Sep 23;12(19):9541. doi:10.3390/app12199541

17. Rodríguez Tembrás V. Two Languages, One Goal: Code-Switching in Doctor–Patient Communication in the Galician Healthcare System. Languages. 2024 Jun 6;9(6):209. doi:10.3390/languages9060209

18. Alkhlaifat E, Yang P, Moustakim M. Code-switching between Arabic and English during Jordanian GP consultations [Internet]. 2020. doi:10.15290/cr.2020.30.3.01

19. Diwan A, Vaideeswaran R, Shah S, Singh A, Raghavan S, Khare S, et al. Multilingual and code-switching ASR challenges for low resource Indian languages. In: Interspeech 2021 [Internet]. 2021 [cited 2026 Mar 9]. p. 2446–50. Available from: http://arxiv.org/abs/2104.00235 doi:10.21437/Interspeech.2021-1339

20. Guo Y, Hu D, Ziqi Chow E, Tam S, Perret D, et al. Clinicians’ Rationale for Editing Ambient AI–Drafted Clinical Notes: Persistent Challenges and Implications for Improvement [Internet]. medRxiv; 2026 [cited 2026 Mar 9]. p. 2026.02.20.26346729. Available from: https://www.medrxiv.org/content/10.64898/2026.02.20.26346729v1 doi:10.64898/2026.02.20.26346729

21. Abridge [Internet]. 2024 [cited 2026 Mar 9]. Record an Encounter in Multiple Languages. Available from: https://support.abridge.com/hc/en-us/articles/30235001206419-Record-an-Encounter-in-Multiple-Languages

22. Pioneering the Science of AI Evaluation [Internet]. [cited 2026 Mar 9]. Available from: https://www.abridge.com/ai/science-ai-evaluation

23. Liu H, Zhang H, Zhang Q, Zhang X, Shi D, Chng ES, et al. Code-switching Speech Recognition Under the Lens: Model-and Data-Centric Perspectives [Internet]. arXiv; 2025 [cited 2026 Mar 10]. Available from: http://arxiv.org/abs/2509.24310 doi:10.48550/arXiv.2509.24310

24. Ugan EY, Pham NQ, Waibel A. DECM: Evaluating Bilingual ASR Performance on a Code-switching/mixing Benchmark. In: Calzolari N, Kan MY, Hoste V, Lenci A, Sakti S, Xue N, editors. Proceedings of the 2024 Joint International Conference on Computational Linguistics, Language Resources and Evaluation (LREC-COLING 2024) [Internet]. Torino, Italia: ELRA and ICCL; 2024 [cited 2026 Mar 10]. p. 4468–75. Available from: https://aclanthology.org/2024.lrec-main.400/

25. Sood A, Srivastava DM. A Review of Automatic Speech Recognition Technology and its Applications in the Medical Field. J Stud Res. 2025 Feb 28;14(1). doi:10.47611/jsrhs.v14i1.8753

26. Quiroz JC, Laranjo L, Kocaballi AB, Berkovsky S, Rezazadegan D, Coiera E. Challenges of developing a digital scribe to reduce clinical documentation burden. NPJ Digit Med. 2019 Nov 22;2:114. doi:10.1038/s41746-019-0190-1 PubMed PMID: 31799422; PubMed Central PMCID: PMC6874666.

27. Kuligowska K, Stanusch M, Koniew M. Challenges of Automatic Speech Recognition for medical interviews - research for Polish language. Procedia Comput Sci. 2023 Jan 1;27th International Conference on Knowledge Based and Intelligent Information and Engineering Sytems (KES 2023)225:1134–41. doi:10.1016/j.procs.2023.10.101

28. Dungavath SN, Nataraj KS, Tiwari N. Self-Training and Error Correction using Large Language Models for Medical Speech Recognition. In: 2024 IEEE Conference on Engineering Informatics (ICEI) [Internet]. 2024 [cited 2026 Mar 10]. p. 1–6. Available from: https://ieeexplore.ieee.org/document/10912300 doi:10.1109/ICEI64305.2024.10912300

